# Notification and recordkeeping of occupational mesothelioma in India

**DOI:** 10.1101/2025.02.11.25322115

**Authors:** Raja Singh, Arthur L Frank

## Abstract

This paper studies the number of occupational mesothelioma cases that have been reported as per law to the factories and mines regulators in India. Zero cases of mesothelioma, which is a notified/notifiable disease under the Mines Act 1952, and notified/notifiable under the broad category of occupational cancers under the Factories, Act 1948 have been notified between 2004 to 2024 (with the factory data compiled only for 8 years). This highlights many issues, including the lack of recordkeeping of occupational diseases when there are cases of mesothelioma being reported by hospitals under the National Cancer Registry Program and many being reported in the scientific literature from India. Though lack of data may not mean the lack of disease, these cases of mesothelioma may either point to a non-occupational exposure to asbestos as an aetiology for such cases, or fixing the recordkeeping and notification of cancers, including mesothelioma, as many occupational cases are reported in the scientific literature. This is especially true as India is one of the largest users and processors of imported asbestos, user of talcum powder and has other potential sources of exposure. With mesothelioma being a surrogate for asbestos exposure, this public health hazard needs focussed implementation of regulated safeguards both occupationally or through non occupational exposures.

## Introduction

The Indian parliament in 2020 passed the Occupational Safety, Health, and Working Conditions Code(1). This legislation subsumed 13 existing major labour laws in India (2). Two important laws that were subsumed are the Factories Act, 1948 and the Mines Act, 1952 (3,4). As of today, in the absence of the Indian Government bringing the Occupational Code 2020 into action, the Factories Act 1948 and the Mines Act 1952 remain enforceable (Further, the provisions of both these legislations have been retained in spirit in the new code). Both these legislations have the provision for notification of occupational diseases. Under Section 89 of the Factories Act, 1948, it is a legal requirement for the manager of the factory or the medical practitioner attending to a worker in the factory, to report to the Chief Inspector of Factories, if any worker has contracted any disease that been listed in the Third Schedule of the Factories Act, 1948. Same is the case with the Mines Act, 1952, in which, Section 25 states that the owner, agent/manager or any medical practitioner attending to a person has to report to Chief Inspector of Mines if any person employed in the mine contracts a ‘disease connected with mining operations’ as notified by the central government. The Central Government, under the Mines Act, 1952 has notified ‘ the cancer of the lung or the stomach or the pleura and the peritoneum (i.e. mesothelioma)’ as a reportable/notifiable disease since 1986. Further, in the case of the Factories, Act, 1948, the ‘Third Schedule’ lists 29 notifiable diseases which includes a cancer of the skin separately and has an entry for ‘Occupational Cancer’ separately. Mesothelioma as a matter of fact is often an occupational cancer, and to record cases of mesothelioma, the occupational cancer list may be updated.

The authors have performed an earlier study looking at the number of mesothelioma cases throughout India (5). The study found 2213 cases of mesothelioma from 2012 onwards till ten years and found 1126 cases in a period of 4 years 2012-2016. The National Cancer Registry Program or NCRP of the Government of India reported 54 cases during the same time period (6). This is because despite being operational since 1981, the registry program has been able to cover only 16% of the Indian population (7,8). In the study as above, it was found that only 21% hospitals in the study were part of the NCRP. Similarly, studies by other researchers in two hospitals in Rajasthan and Gujarat have reported 76 cases (2015-2020) and 126 cases (2015-2019) of mesothelioma respectively (9,10). It was noted that in the first study, all patients denied any past exposure to asbestos, but 91% patients had a history of mining, direct or indirect, in marble and granite mines (or quarries) (9). This also means that some of these are likely occupational mesotheliomas, which should have been reported under the Mines Act, 1952. Even if this link to occupational aetiology cannot be firmly established, it does become a case and should be either reported as occupational, and if not to be able to ascertain, whether the cases may be non-occupational. There are other studies where factory workers have been exposed to asbestos and this has been reported as the probable cause of mesothelioma. The data from the study of Indian mesothelioma numbers is unclear on whether these cases are from occupational or non-occupational exposure. (11). This study aims to discuss this gap as the number of occupational mesotheliomas, or occupational diseases in general have not been compiled before and reported as required. It is vital to understand this to inform policy and to allocate already legislated responsibility to various stakeholders in the management of occupational diseases in India. Further, knowing mesothelioma case numbers from occupational settings will specifically inform about the causative factor asbestos exposure and will help ascertain whether the source is occupational and/or non-occupational. This is important as asbestos is still widely imported in India, used in India, processed in Indian factories, used by Indian population and disposed of into the general environment (12–15). There are also identified cases of other minerals which have asbestos as a contaminant to which the Indian population is exposed to in occupational and non-occupational settings, without awareness or precautions (16–19).

Further, both statutes have a provision where the medical practitioner can be fined if they fail to notify occupational diseases under the Section of Factories Act, 1948 and under the Section of the Mines Act, 1952.

### Aim

To study the number of occupational mesothelioma cases that have been recorded and notified in India.

Specific Objectives are firstly, to assess the number of mesothelioma cases that have been recorded in a time period (20 years) by the Chief Inspector of Mines as specified under the Mines Act, 1952. Secondly, to assess the number of mesothelioma cases (recorded with other occupational cancers except a form of skin cancer which is recorded separately) under the Factories Act, 1948. There is a third objective, which is to report any incidental findings that were derived in the process of achieving the above objectives.

## Methodology

In India, under the Mines Act, 1952, the Chief Inspector of Mines is the designated authority to which mine management from across the country must report and notify occupational diseases. The Directorate General of Mines Safety, Dhanbad has been designated by the Government of India as the Chief Inspector under the Mines Act, 1952 (20). An information request under the Right to Information Act, 2005 was filed to request data as DGMS is a public authority. The same process of requesting information was done from the Directorate General of Factory Advice Services and Labour Institutes, or DGFASLI, which is a Government of India organisation under the Ministry of Labour and Employment. The period of Information was from 2004 to 2024.

Information was directly provided by the DGMS in a compiled form and the same has been reported below.

The DGFASLI stated that the report is compiled by state/Union Territory (UT). This is done by its internal statistics cell from the Chief Inspector of Factories/Directorate of Industrial Safety and Health of individual states and is published annually in the form of a Standard Reference Note report, which is published by DGFASLI on its website (21). These reports were individually analysed and it was found that the reports were available on the website only from 2006 to 2023 which means the data was available for the years 2005-2023. Further, the compilation of occupational diseases was only made from the year 2016 onwards, which means the data is only available from 2015 until 2022. The available date was compiled and has been presented below. It must be noted that there is a difference between the factories regulation and the mining regulation. The primary difference is that factory safety falls under the purview of the state governments and the Central government provides guidance through the DGFASLI, but the mine safety is directly under the purview of the Indian central government. This is despite the fact that land revenue and mining revenue being under each state, while safety is under the centre.

It should be further noted that the information released under the Right to Information Act, 2005 is information in the public domain. Further, it implies that the information is non personal and does not have any human identifier as public authorities by law can only release non personal information (22). The study involves no human participants and required no ethics approval.

## Results

In the time period between 2004 to 2024, the DGMS stated that, no single case of mesothelioma were reported to the Chief Inspector of Mines from throughout India.

Further, the data from 2015 to 2022, from the DGFASLI, also had no cases of occupational cancer (as mesothelioma is not directly classified under the Factories Act, 1948) reported state-wise/UT-wise by the Chief Inspector of Factories to the DGFASLI. It is further noted that there were many states such as Uttar Pradesh, West Bengal and Punjab which at one or more instances failed to provide information to the DGFASLI on the number of cases of occupational diseases that has been notified to the state Chief Inspector of Factories.

Some incidental results were reported. First, the DGMS reported that no medical practitioner has ever been fined for not reporting occupational diseases.

Further, the diseases that have been reported over the years under the Factories Act, 1948 include silicosis, noise induced hearing loss, silico-tuberculosis, byssinosis, irritant contact dermatitis, toxic jaundice and pneumoconiosis. The states that have consistently reported are Gujarat and Maharashtra.

As far as the reporting of notified/notification of diseases is concerned under the Mines Act, 1952, the diseases that have been reported, by the states that did report are silicosis, coal worker’s pneumoconiosis and noise induced hearing loss. No case of manganese poisoning (nervous type), asbestosis, mesothelioma, contact dermatitis (caused by direct contact with chemicals) and pathological manifestations due to radium or radioactive substances have been reported to the Chief Inspector of Mines from 2004 to 2024.

## Discussion

As far as mesothelioma in India, there have been research papers from academic institutions that have reported cases (5,9–11,23–25). There are other cases, where research papers have been published, but the medical records section of the same hospital deny any case within that period (24,26). It is concerning that mesotheliomas have been diagnosed and reported at large hospitals, but these are not necessarily reported to the factory or the mine regulators.

There are reported mesothelioma cases in the country that have been associated with occupational exposure from research papers published in India, but have not been reported to regulators as from Rajasthan where out of 76 cases, 69 persons getting the disease had all come from the same demographic region and had a direct or indirect exposure to mining and quarrying industry of marble and granite (9). This lack of reporting, at the simplest, may be considered a violation of the law, but it also points at a serious systematic issue, with respect to creation of a mechanism for doctors to report occupational cases, and reflects the general issue of lack of recordkeeping of diseases in India (27). These reported cases, or diagnosed-but-not-reported cases likely do not tell the complete pictures as there may be cases which may never reach the hospital, or which are mis-diagnosed or which are not recorded internally in hospital clinics. As a country, if India does not measure disease incidence, then there will be a serious problem in the management and prevention of such diseases . This is all the more true when there are some occupational diseases which overlap with other serious infectious diseases which India plans to eliminate, like tuberculosis, which overlaps with silicosis causing silico-tuberculosis, which is primarily seen in occupational settings (28). Taking occupational health seriously, by first starting to note the accurate count of cases, will enable reduction of a preventable burden disease (29). It will also make economic sense as the disease burden can in the future bring down the value generated by the workmen and employees (30).

It must also be noted that regulation of labour in mines in India is the duty of the Central Government as it is part of the Union list of the Constitution of India (31). Almost all industries are part of the state list and the welfare of labour under the concurrent list of the Indian Constitution. This means that for industries, the Central Government can mostly play an advisory role, and for labour regulation, both the Centre and State must agree on the same rules. For this reason, the data of notification of disease from mines is from the Central Government controlled Directorate General of Mines Safety, whereas the data on implementation on the factories regulation should be derived from the Chief Inspector of Factories from the states. The data collected in this paper from the DGFASLI is data collected in DGFASLI’s advisory role. Data collected from the Chief Inspector of States respectively would be the most reliable data. This data collection from the Chief Inspector of the states directly has not been done in this present study. This is a limitation and there is scope for future work with data collected from the states directly. It must also be noted that the various laws including the Mines Act, 1952 and the Factories Act, 1948 are in the process of merging into a set of four labour codes, with one specifically dealing with occupational health and safety. It will require serious political and administrative will as the codes largely have components from the older laws, which too had the best of intentions, but their compliance may not be where they should at this time.

Apart from the mechanism of recording and notification of occupational mesotheliomas (through Mines and Factory Regulator), there is also the National Cancer Registry Program or NCRP, on which the authors have published a previous study (5). Cancers, including mesotheliomas, are underreported in India since Indian cancer recording is poor and covers only about 16 % of the total population (32). The authors in the previous study found that in 83 hospitals from which cases were collected, only 21% were under the registry program directly, and 1126 cases of mesothelioma were recorded for one four year period, for which the cancer registry program reported only 54. In another study from a cancer hospital in Ahmedabad, 62 cases of mesothelioma were reported from a hospital in Gujarat in which usually, as reported by another study, about 1/3^rd^ of cases may be reported from the State of Rajasthan itself (5,10). Discussion about cancer notification in India is being opposed by the Indian Health Ministry despite recommendations by Indian Council of Medical Research-National Centre for Disease Informatics and Research or ICMR-NCDIR. ICMR-NCDIR which is a government research institute running the NCRP, due to a lack legal mandate to report cancer cases may be unable to enable full recordkeeping of cancer (7).

There is also the issue of defining of the jurisdiction of what is an occupational disease in a factory versus what may be an occupational disease in a mine. A mine involves extraction from the ground, whereas further processing and milling of minerals may be in the jurisdiction of factories (16). This requires more thought and in the process of jurisdiction division, the true case incidence should be reported and recorded. Silicosis has been reportable under the factories law as well as the mines law, which indicates it is not just be a mining issue, but linked to additional processing. Similarly, the case of mesothelioma which is reportable as such under the mining law and is reportable as occupational cancer under the factories law.

Mesothelioma can be considered as a surrogate for exposure to asbestos (33). Mesothelioma in particular, a focus of this study, is of grave concern as India continues to be a major importer, processor and user of this carcinogen that has been banned completely in some 68 countries across the world (12,34,35). In 1986 the Indian government stopped asbestos mining and similar order was issued in 1993 (36). Despite the mining ban, India has become the largest importers of asbestos. Data derived from the Directorate General of Commercial Intelligence and Statistics, Ministry of Commerce and Industry, Govt. of India, in the period 2021-22 to 2023-24 (Updated till Jan 2024), shows the three year average import of asbestos (chrysotile asbestos with HSN number 2524) has been from Russia (44.40% or Rupees 929.90 crore), Brazil (36.63% or worth 767.10 crore rupees), and Kazakhstan (18.26% or 382.39 crore rupees) (37). There are other places with minor import amounts including China, Poland, Georgia, Vietnam, Turkey, etc. Brazil, another developing country, has banned the use of asbestos within the country, but still is reported to export to India (38). These import sources replaced Canada after its mining ceased (39). In countries where the use of asbestos has ceased, there has been a decrease of mesothelioma cases (40) . In India, even after a mining ban, the government, except for the Environment Ministry, has mostly seems to defend the import, processing and continued use of asbestos (12). There has been an effort by a parliamentarian to introduce a private member’s bill in the Indian parliament to ‘provide for a total ban on use and import of white asbestos in the country’ in 2014 (41). Such bills serve as a statement of urgent need to highlight an important issue like this, despite these not getting passed as they are not government initiated bills (42). In the Rotterdam Convention, having a list of hazardous chemicals and pesticides, chrysotile has not been included and the Indian government supports this exclusion, while in the Factories Act, 1948, asbestos is listed in the First Schedule which lists ‘industries involving hazardous processes’ (3,12,43). Even in environmental clearance, all projects involving ‘asbestos milling and asbestos based products’ are in category ‘A’ along with mining, oil drilling, large river valley and thermal projects. Category ‘A’ projects can only be approved by the Indian Central Government on recommendations of an Expert Appraisal Committee and require mandatory clearance and public consultation. These categories are based on ‘the spatial extent of potential impacts and potential impacts on human health and natural and man-made resources’ (44).

All forms of asbestos, including chrysotile, are carcinogenic and a threat to human health (45). As a carcinogen, the World Health Organisation states ‘…There is no safe level of asbestos exposure (46).’ This is specifically true for all carcinogens, including asbestos, and the exposure is not limited to the dose (47). Inhaling asbestos fibres can lead to scarring of the lung, which can result in loss of lung function, disability & death (48).. Asbestos is a risk factor for developing disability & deadly lung diseases decades after exposure, which can range from 15 years to 40 years or more.

A study from Mumbai stated that in the near future, ‘there will be at least 12.5 million asbestos-related disease patients and 1.25 million asbestos related cancer patients worldwide, and half of these will be in India (49).’ Mesothelioma, which takes decades to manifest, is also predicted to occur in 15% of all workers related to ship breaking activities where asbestos containing materials are handled (50). India’s Ministry of Environment, Forests and Climate Change in its ‘Vision Statement on Environment and Human Health’ stated that ‘alternatives to asbestos may be used to the extent possible and use of asbestos may be phased out (51).’ This vision is in line with the position of the International Labour Organisation which passed a resolution in the 95th Session in 2006, where it called for ‘elimination of the future use of asbestos and identification and proper management of asbestos currently in place as most effective means to protect workers from asbestos exposure and to prevent future asbestos related deaths’ (52). It further stated that the 1986 resolution, which considered regulated use of asbestos, without eliminating its use, ‘should not be used to provide a justification for, or endorsement of, the continued use of asbestos.’ India has not ratified the two conventions related to asbestos and allied disease issues namely the Asbestos Convention, 1986 and the Occupational Cancer Convention, 1974 (53,54).

India’s Supreme Court has dealt with the occupational use of asbestos through two judgements. The first one being 1995 Consumer Education & Research Centre & Ors v. Union of India & Ors. judgment or the CERC 2005 judgement and second one being the Kalyaneshwari vs Union of India, 2011 judgment or the Kalyaneshwari 2011 judgement (55,56). The compliance status of these two judgments is not within the scope of this paper, but there may be much remaining scope for full compliance of the judgements and their intent.

Indian government agencies have also undertaken various studies related to the occupational use of asbestos. This list includes the 2019 study by the Directorate General Factory Advice Service and Labour Institutes or DGFASLI, under the Ministry of Labour and Employment, Government of India was titled ‘National Study of Occupational Safety, Health and Working Environment in Asbestos-Cement Product Industries, 2019’ (57). The authors investigated the study from information provided by DGFASLI (58). The study did not include the investigation of any mesothelioma cases, and these details were not available in the report. The first author also sought details about the coverage of malignancy with long latency periods from 15-40 years, such as mesotheliomas, to which the DGFASLI stated to not have information in this report. DGFASLI also stated they had no information in the report regarding any retired workers, as they were not covered in the study. These retired workers, if also investigated, would have provided more insight to long latency malignancies like mesotheliomas much after their actual exposure in workplaces. The study may have covered some relevant areas of investigation, but surely lacks in factoring in incidence of mesotheliomas and investigating long term latency of disease that is caused by exposure to asbestos. This exclusion in spirit is not in line with the order of the Supreme Court of India which had directed all asbestos industries to ‘maintain and keep maintaining the health record of every worker’ up to a minimum period of 40 years from the beginning of the employment. It is also possible that industries may simply keep healthy workers and fire the unhealthy ones (or coerce them to resign) leading to a “healthy worker effect”[13]. The second study has been by the National Institute of Occupational Health or NIOH and there has been another one by the Indian Bureau of Mines or IBM (12). Future work can look into these studies in detail.

Asbestos milling and asbestos products industries must get approval from the Central Government’s Ministry of Environment, Forest and Climate Change in the form of an Environmental Clearance which is granted after performing an Environmental Impact Assessment (59). These steps include compliance with the Kalyaneshwari judgement (56). Further studies can look into the compliance of Environmental Clearance requirements by asbestos industries.

Mesothelioma, a ‘disdained member of thoracic oncology’ must be better studied to improve overall management and also this disease should be prevented by strict regulation or even stopping the use of asbestos (25). There is no safe level for carcinogens, including asbestos which causes mesothelioma (47). It is important that apart from occupational exposures, the whole life cycle of asbestos including non-occupational exposures must also be considered, including installation, use, demolition and disposal of its products (12,60). Care must also be taken in abandoned asbestos mines which after the ban on mining remain unremedied (61).

## Conclusion

The study aimed to find the number of occupational mesotheliomas that have been notified and recorded in India. This means the number of mesothelioma cases that have been recorded by the Chief Inspectors of Factories under the Factories Act, 1948 and the Chief Inspector of Mines under the Mines Act, 1952. The results show that zero cases have been notified and recorded. This can be interpreted to either mean that the mesotheliomas recorded in India by other studies are completely from non-occupational exposure to asbestos, or secondly, despite occupational mesotheliomas suggested by research papers, no reporting to the factory and mine regulators means that there is an absence of recordkeeping and a violation of the provision of the two laws (Factories Act, 1948 and Mines Act, 1952) made by the Indian Parliament.

### Recommendations

It is recommended that, firstly, the provisions in both the laws where the medical practitioners are penalised on failure to report occupational diseases must be strictly implemented. Non-reporting by doctors itself despite their moral and legal responsibility could be avoided by enforcement of fines. Medical practitioners must be made aware during their education and during their practice of their duty to notify occupational disease cases. The National Medical Commission may take up this cause and create a mechanisms to make doctors aware of their responsibility to report occupational diseases. Secondly, the Chief Inspector of Mines at the Central level and the Chief Inspector of Factories at the state level must create infrastructure and mechanisms so that doctors are constantly reminded of their duty and there should be ease of reporting. There should be regular outreach programs at all hospitals, especially tertiary care facilities and referral centres by these authorities. Thirdly, the possibility of conflict of interest at the level of states where factories and mines are wealth generators and may not be regulated in spirit for being revenue creators, must be re-evaluated from the social and financial cost of an unhealthy workforce. The healthcare cost due to occupational diseases must be quantified and this must be subtracted from the sum total of revenue generated from factories and mines. Fourthly, there should be coordination between the National Cancer Registry Program run by the Indian Council of Medical Research-National Centre for Disease Informatics Research and the recordkeeping done by the mining and factory regulators.Instances where there are cases recorded within an area of a cancer like mesothelioma by NCRP but not by the mine regulator decreases confidence on laws and their enforcement should be avoided. Fifthly, cancer must be made notifiable across the country, despite it being a non-communicable disease. 17 states in India have notified cancer despite the centre refusing to do in line with ICMR-NCDIR recommendations (7). This will enable universality in reporting and prevent underreporting especially in cases where there is no clear-cut differentiation between occupational and non-occupational aetiology can also be recorded. Sixth, mesothelioma has been a separately classified occupational disease under the Mines Act, 1952 since the notification in 1986, but after the promulgation of the Occupational Safety, Health and Working Conditions Code, 2020, it has not been retained in the list of notifiable diseases in the Third Schedule. While other miner’s related diseases such as coal miner’s pneumoconiosis, seem to be retained, mesothelioma is absent in the new list. It seems to have been merged like in the Factories Act, 1948 with Occupational Cancer. It is recommended that being a rare malignancy, associated with a specific exposure to asbestos, it should be listed as a separate entry in the Third Schedule of the new Occupational Safety, Health and Working Conditions Code, 2020. Seventh, studies, including the one done by DGFASLI in 2019, may have missed the component of latency in causation of mesothelioma, as past records may not have been evaluated. It should be pertinent that future studies related to asbestos exposure and mesothelioma may have the following components: a. Consideration of long latency of disease caused by asbestos exposure, which may be 10/15 to 40 years or more. B. Inclusion of sampling of the environmental conditions, non-occupational exposures and para occupational exposures in studies performed related to asbestos exposure. C. Consideration that permissible levels may not exist for carcinogenicity.

Eighth, in compliance of the CERC 1995 judgment and the Kalyaneshwari 2011 judgment of the Supreme Court of India, the directions which may not have been complied with, may be complied with as non-compliance is a contempt of the orders of the Hon’ble Court. Ninth, The International Labour Organisation’s two conventions, one being the Occupational Cancer Convention, 1974 and the other being Asbestos Convention, 1986, which remain non-ratified in India, should be reconsidered for ratification (53,54,62). Tenth, history taking by doctors in mesothelioma suspected cases must not only take an occupational history, but also a history of other factors related to exposure such cosmetic talcum powder, exposure to other minerals with asbestos contamination, proximity to asbestos an factory or mine, or use of asbestos products, para-occupational exposure, some of which may be known and be recallable and others may require some further questioning by the history taking physicians (16,18,19,63,64). Eleventh, there must be focus on worker health and safety. By a recent amendment to the Mines and Minerals (Development and Regulation) Act, 1957, there has been the creation of the District Mineral Foundation (65). This foundation with a decentralised mandate should concentrate on the health and safety of workers by provision of Personal Protective equipment, regular health check-ups and all other mechanisms for the well-being of workers. As per the latest National Mineral Policy of India, DGMS ‘should be further strengthened’ so that the miners’ health and mine safety can be ensured (66). The *Pradhan Mantri Khanij Kshetra Kalyan Yojana* (PMKKKY) translated as Prime Minister’s Mining Area Welfare Scheme with its inclusion of Health care as a priority area for very localised and contextual solutions must be followed both in letter and spirit, and it should not only emphasise health infrastructure development, but provide full provisions for effective operations and maintenance, including proper staffing (67). Lastly, apart from focus on recordkeeping and history-taking, a general increased interest in occupational medicine must be focussed in India.

## Data Availability

All data in the present work are contained in the manuscript

## Future Work

Future work in this area may focus on getting data from the state Chief Inspector of Factories from all state wises (and UTs) including number of medical practitioners that have been penalised for not reporting occupational diseases (including mesothelioma). There is also need to check compliance with the Factories Act, 1948 with respect to the appointment and facilitating the work of certifying surgeons. There is a need for performing epidemiological studies related to mesothelioma. This means that long term prospective studies or very thorough case control studies must be performed to ascertain hotspots in terms of locations, exposures and possible unknown factors. Doing regular check-ups as stated in the law for miners in mineral mines containing asbestos contamination can serve as a ready source of data for such studies. Identification of Indian materials, consumer products, other minerals (like marble, soapstone, etc), industrial products or other sources containing asbestos that have been available to the public in the past decades must be performed and such exposures listed as these may serve as a guide to doctors performing history taking from mesothelioma patients. Further work can also check the compliance status of the judgments of the Supreme Court of India as well as the environmental clearance requirements by asbestos industries. A study looking into the three important studies by Indian agencies can also be performed.

## Declarations

No external funding has been received for this work. The first author declares no conflict of interest. The second author conducts medical–legal work regarding asbestos, primarily for plaintiffs. The study uses data available in the public domain, involves no human participant, involves no animals or tissue and hence requires no ethical clearance. All data in the present work are contained in the manuscript.

## CRediT Author Contributions

Conceptualization: RS; Data Curation: RS; Investigation: RS; Methodology: RS; Project Administration: ALF, RS; Resources: ALF, RS; Supervision: ALF; Validation: ALF; Writing-original draft: RS; Writing-review & editing: ALF, RS;

